# Contact tracing and isolation of asymptomatic spreaders to successfully control the COVID-19 epidemic among healthcare workers in Milan (Italy)

**DOI:** 10.1101/2020.05.03.20082818

**Authors:** Stefan Mandić-Rajčević, Federica Masci, Eleonora Crespi, Sara Franchetti, Anna Longo, Ilaria Bollina, Serena Velocci, Alessandro Amorosi, Riccardo Baldelli, Luisa Boselli, Lucia Negroni, Alessandro Zà, Nicola Vincenzo Orfeo, Giuseppe Ortisi, Claudio Colosio

## Abstract

**Objective:** To study the source, symptoms, and duration of infection, preventive measures, contact tracing and their effects on SARS-CoV-2 epidemic among healthcare workers (HCW) in 2 large hospitals and 40 external healthcare services in Milan (Italy) to propose effective measures to control the COVID-19 epidemic among healthcare workers.

**Design:** Epidemiological observational study.

**Setting:** Two large hospitals and 40 territorial healthcare units, with a total of 5700 workers.

**Participants:** 143 HCWs with a SARS-CoV-2 positive nasopharyngeal (NF) swab in a population made of 5,700 HCWs.

**Main outcome measures:** Clinical data on the history of exposure, contacts inside and outside of the hospital, NF swab dates and results. A daily online self-reported case report form consisting of the morning and evening body temperature and 11 other symptoms (cough, dyspnoea, discomfort, muscle pain, headache, sore throat, vomiting, diarrhoea, anosmia, dysgeusia, conjunctival hyperaemia).

**Results:** Most workers were tested and found positive due to a close contact with a positive colleague (49%), followed by worker-initiated testing due to symptoms (and unknown contact, 28%), and a SARS-CoV-2 positive member of the family (9.8%). 10% of NF swabs performed in the framework of contact tracing resulted positive, compared to only 2.6% through random testing. The first (index) case caused a cluster of 7 positive HCWs discovered through contact tracing and testing of 250 asymptomatic HCWs. HCWs rarely reported symptoms of a respiratory infection, and up to 90% were asymptomatic or with mild symptoms in the days surrounding the positive NF swab. During the 15-day follow-up period, up to 40% of HCWs reported anosmia and dysgeusia/ageusia as moderate or heavy, more frequently than any other symptom. The time necessary for 95% of HCWs to be considered cured (between the positive and two negative NF swabs) was 30 days.

**Conclusion:** HCWs represent the main source of infection in healthcare institutions, 90% are asymptomatic or with symptoms not common in a respiratory infection. The time needed to overcome the infection in 95% of workers was 30 days. Contact tracing allows identifying asymptomatic workers which would spread SARS-CoV-2 in the hospital and is a more successful strategy than random testing.

**What is already known on this topic?:** There are more than 3 million SARS-CoV-2 positive cases and more than 200,000 deaths attributed to coronavirus disease (COVID-19) worldwide.

Commonly reported symptoms of COVID-19 include fever, cough, dyspnea, sore throat, muscle pain, discomfort, and many prevention strategies are based on identifying these symptoms of infection.

The virus can be spread even by asymptomatic patients or patients with mild symptoms, and healthcare workers (HCWs) represent 10% of overall cases and often more than 10% of hospital personnel are commonly infected.

HCWs represent both a vulnerable population and an irreplaceable resource in the fight against this epidemic and further analysis is needed to show how and why they get infected and introduce successful prevention measures.

**What this study adds?:** The first (index) case in our study was infected by a family member, but due to close contacts with colleagues managed to infect other 7 HCWs. Contrary to a common expectation that HCWs get infected from patients, they regularly get infected by other HCWs.

Up to 90% of HCWs were asymptomatic or had only mild symptoms. Random testing for SARS-CoV-2 was not efficient. Active search for suspect cases through contact tracing is the strategy of choice to identify most of the positive HCWs.

Most HCWs remained asymptomatic during the 15-day follow-up period, and even in the days prior to the positive NF swab. Anosmia and ageusia/dysgeusia were reported more commonly than classic symptoms of a respiratory infection.

Contrary to the recommended quarantine of 14 days, 30 days were necessary for 95% of the workers to be declared cured (two negative NF swabs)

## Introduction

On the 30th of January 2020, the World Health Organization (WHO) declared the outbreak of the novel coronavirus disease 2019 (COVID-19) as a Public Health Emergency of International Concern. At the time of the writing of this paper, almost 3 million cases and almost 200,000 deaths have been reported worldwide (1). Most prominent symptoms include fever, dry cough, headache, sore throat and sneezing, although a growing number of reports underline asymptomatic and patients with mild symptoms having the same viral load as symptomatic patients and spreading the infection in the general population and among healthcare workers (HCW) (2–5). Most published reports on COVID-19 patients underline that HCWs get infected regularly and they represent one of the most vulnerable groups during this pandemic (6,7). The safety of HCWs is key not only to fight this international biological threat through their care for the critically ill patients, but also to prevent them from transmitting the virus.

WHO and European Centre for Disease Control (ECDC) recommendations for the rational use of personal protective equipment agree on the use of: a) a medical (surgical) mask, eye protection, long-sleeved water-resistant gown, gloves, and keeping 1 m distance when dealing with suspected or confirmed COVID-19 cases; b) a respirator (N95 or FFP3), eye protection, water-resistant gown and gloves during aerosol generating procedures; c) a medical (surgical) mask to be worn by suspected or confirmed COVID-19 patients, or any patient with respiratory symptoms. Good hand hygiene should be kept using 70% alcohol based disinfectants or soap and water (washing at least 20 seconds) (8–10). WHO definitions of suspect cases require at least mild symptoms, although contact tracing and adjustment of suspect case definition is encouraged (11). Inclusion of contact tracing and testing of asymptomatic patients can help identify potential spreaders of SARS-CoV-2 in hospitals, but with a high logistic burden (12). Having in mind that around 10% of infected persons are HCWs, and that most hospitals report 10% of staff getting infected (13), the additional burden might be worth it. Additional challenges are posed by the removal from workplace and return-to-work procedures which depend on the expected duration of the disease and a negative nasopharyngeal swab, which can be false negative (14).

In this frame, we developed our own experience in the Territorial Socio-Sanitary public company of the Saints Paolo and Carlo of Milano, Italy (TSSC). The first SARS-CoV-2 infected HCW in our structure was confirmed on February 27^th^, 2020. Since that moment, we applied a protocol based on five main steps:

1. To identify a case (or cases) which could be diagnosed by any of our hospitals or reported by any other affordable source (patients and workers);
2. To identify symptomatic workers and to verify the existence of an infection;
3. To conduct an internal epidemiological survey addressed at identifying all close contacts of the infected subjects;
4. To “biologically” isolate these contacts, in the hospital and in the private life, and immediately perform a nasopharyngeal swab;
5. To decide, based on clinical and laboratory data, the return-to-work policy safe for the HCWs and their colleagues.

**The aims of this paper are to analyse the effects of preventive measures taken in our TSSC to tackle the epidemic, the onset and follow-up of COVID-19 symptoms in the typical working age population of HCWs, the sources of infection in HCWs and the duration of the infection to propose adequate, evidence-based, and effective measures to control the COVID-19 epidemic in healthcare institutions and establish an appropriate quarantine period**.

## Material and Methods

### The setting

Our TSSC is composed of two public hospitals of Milan and 40 territorial healthcare institutions (providing various services such as vaccination, preventive care, administration), which are part of the Region of Lombardy (Northern Italy) public healthcare system. The two hospitals employ a total of 4142 workers (healthcare workers and non-healthcare workers in a healthcare setting, all referred to as HCWs in our report). There are around 1500 workers in 40 territorial healthcare institutions. Out of 5,700 workers, 70% are female, with a mean age of 46 years. It is estimated that around 150,000 patients are examined in our emergency rooms of the two hospitals every year, with an average of 50 admissions per day in around 800 hospital beds available. The hospitals provide all diagnostic and medical procedures covering most existing medical specialties. San Paolo hospital is also the home of the Department of Health Sciences of the University of Milano, which integrates it into the University of Milan academic system. Workers of the whole groups of structures are provided with occupational health services in the healthcare setting.

Since at the beginning of the epidemic, most administrative workers started smart working, with only a few exceptions. During our routine occupational health surveillance of workers, we transferred the most vulnerable subjects (the elderly, those affected by chronic diseases) from the most risky departments (i.e. emergency room) to “safer” departments. Most vulnerable workers got the possibility to remain home, without any impact on their earnings. Our study is addressed at the health care personnel who continued their activities during the crisis.

### Ethical approval

All data presented in this paper were extracted from the health surveillance files, and no experimental activity has been carried out. Nevertheless, all our workers are informed about our health surveillance procedures and have signed an informed consent regarding the data collection and analysis. No ethical approval was deemed necessary by the Ethical Committee of the Saints Paolo and Carlo Hospitals.

It was not appropriate or possible to involve patients or the public in the design, or conduct, or reporting, or dissemination plans of our research

### Management of HCWs in the frame of SARS-CoV-2 pandemic

***Figure 1*** outlines the procedure and treatment of close contacts, quarantine and return-to-work procedure.

**Figure 1.**
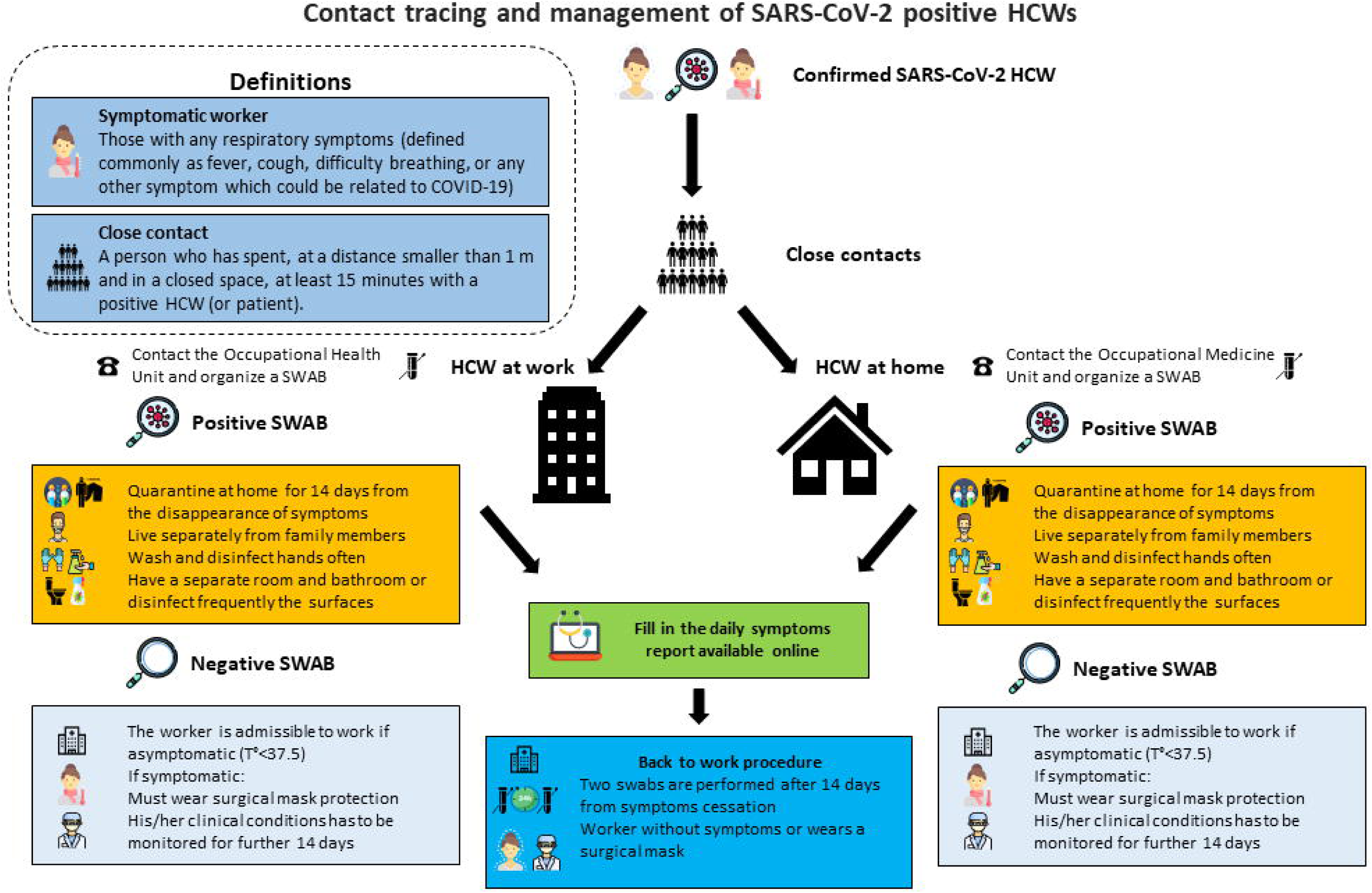
Outline of the HCW procedure for case identification, personal protection, removal and return to work procedure.

A “close contact” was defined as a person who had a face-to-face dialogue or who spent at least 15 minutes in an indoor environment with a COVID-19 patient, without wearing a personal protective device (PPD, e.g. surgical mask). The epidemiological survey was performed by the Health Care Management of the structure and close contacts were reported to our Occupational Health Unit. Symptomatic workers were those with any respiratory symptoms (defined at the beginning as fever, sore throat, cough, difficulty breathing, or diarrhoea). Asymptomatic workers were asked to adopt “source control” and isolation measures to reduce the viral charge and risk of infecting another colleague or a family member. Viral charge reduction or “source control” was performed by asking the worker, even if asymptomatic, to wear a surgical mask both while working, traveling and in the private life. The worker was to take meals separately from the family, live in a separate room and use a dedicated bathroom or at least carefully wash the only bathroom available after use.

Each close contact was required to do a NF swab. Workers absent from work due to respiratory symptoms were asked to come to the hospital for a NF swab when their symptoms would allow it (fever below 37.5°C and other symptoms not preventing to travel). If the swab was positive, the affected worker was placed in mandatory quarantine for a 14-day period. For symptomatic workers the quarantine period lasted for at least 14 days from the full termination of all symptoms, even if the symptoms were reported later during the disease (after the swab). Workers with symptoms were asked to immediately leave the workplace and to go home, adopting the same rules of source control and isolation given to the asymptomatic workers. In case of a negative NF swab, the worker would come back to work after the cessation of symptoms. To reduce the potential impact of false negative NF swabs, all close contacts, even if the swab resulted negative, maintained precautionary source control measures (surgical mask) for a 14-day period after the contact.

During their absence or isolation (negative NF swab but confirmed close contact) all workers were asked to fill in and submit a daily symptoms report which was collected by phone and in paper for the first 5 workers, and then transformed into an online questionnaire to allow an overview and tracking of the whole population of HCWs under observation. This collection was performed in order to point out any sign or symptom indicative of a possible worsening of the health conditions of the worker.

### Return-to-work procedure

We set up a specific procedure to manage safe return to work of the workers absent for COVID-19 or suspected of COVID-19. In particular, a symptomatic COVID-19 worker was considered cured 14 days after the resolution of respiratory infection symptoms and two consecutive negative tests for SARS-CoV-2 at least at 24 hour distance one from the other (15). The definition of “clearance” of the virus indicates the disappearance of detectable SARS-CoV-2 RNA in nasopharyngeal swabs. In case of a positive swab after the 14-day period, another 7 days was added to the quarantine, and then the two swabs were repeated.

We faced an additional problem of readmitting to work HCWs who were absent from work but did not undergo a NF swab during this period. If the worker was absent for 14 days or more (same as COVID-19 positive workers) and/or showed typical symptoms, she or he was treated as a COVID-19 patient. If the period of absence was shorter than 14 days, or the presence of typical symptoms was not recorded, the worker was treated as “close contacts” (followed the “source control” procedure, underwent a NF swab, and symptoms were followed).

### Random testing of workers

Since the 25^th^ of March, a new regulation recommended random testing of HCWs (16). We also report the results of HCWs found positive using this approach.

### Symptoms report

Our study presents the results and analysis of 143 HCWs with a positive NF swab who filled the online symptoms report in the period from the 9^th^ of March until the 8^th^ of April 2020.

HCWs with a positive NF swab, symptomatic HCWs, as well as those under surveillance for a close contact with a positive HCW or a family member were asked to fill in a daily symptoms report. The symptoms report has evolved to improve tracking and adjust for clinical findings during the period in four phases. These phases are described shortly to better explain what data was collected and when:

1. Initially, from the 27^th^ of February until the 8^th^ of March, the daily symptoms report was in paper form and was given to (a few initial) HCWs to fill and submit in writing after the quarantine;
2. From the 9^th^ of March, the symptoms report was put online, and all workers were given a link to an electronic sheet where they would fill and submit the daily report, and their data became available right away. The report included the morning and evening body temperature and there was a free- text field available to add any other symptoms;
3. From the 12^th^ of March, the free-text field for symptoms was converted into specific questions regarding the symptoms (cough, dyspnoea, sore throat, headache, muscle pain, discomfort, vomiting), based on the current knowledge of COVID-19 symptoms;
4. Finally, based on clinical experience, additional symptoms (anosmia, dysgeusia, and conjunctival hyperaemia) were added.

### Nasopharyngeal swab

Nasopharyngeal specimen is the gold standard for swab-based SARS-CoV-2 testing. Nasopharyngeal swab was performed by inserting the flexible wire shaft minitip swab through the nares parallel to the palate until resistance was encountered or the distance is equivalent to that from the ear to the nostril of the patient, indicating contact with the nasopharynx. Then the person performing the swab would gently rub and roll the swab and leave it in place for several seconds to absorb secretions. The swab was then slowly removed while rotating it. All HCWs in charge of taking this sample did a standardized course which allowed all samples to be taken in the same way.

### Nasopharyngeal specimen analysis

Real-time reverse transcription–polymerase chain reaction (rRT-PCR) assay is currently the most reliable and the only available direct method to detect SARS-CoV-2 virus; it is the gold standard method for laboratory diagnosis of COVID19 (17,18).

SARS-CoV-2 is a large positive-sense single-stranded ribonucleic acid (RNA) virus that comprises of four structural proteins, i.e., nucleocapsid protein (NP) that holds the viral RNA, spike protein (SP), envelope protein (EP), and membrane protein (MP), that create the viral envelope. RNA-dependent RNA polymerase gene (RdRp), envelope (E), and nucleocapsid (N) have become key diagnostic targets for SARS-CoV-2 identification.

In the Clinical Laboratory of the Department of Diagnostic Sciences of the San Paolo hospital the NF swab analysis was performed with two kits: GeneFinderTM COVID-19 PLUS RealAmp Kit by ELITech Group and Roche Modular Wuhan CoV N, RdRP and E gene Kit.

Test was Positive if RdRp and E or N or both genes were detected; repeated to confirm Positive if only RdRP or N were detected.

### Statistical analyses

We present a case series of 143 out of 185 SARS-CoV-2 positive HCWs in the study period. Each HCW was connected to their daily reports. The reports were centred around the date of the positive swab, making that day’s report the follow-up day 0 (zero). Days following the positive swab are denoted with positive integers (from 1 to 15), and days leading to the positive swab are denoted with negative integers (from -5 to -1).

The morning and evening temperature were collected as numeric variables. The absence or intensity of each symptom was collected as: absent, light, moderate, and heavy. Categorical variables (i.e. symptom intensity) are presented in tables as the absolute count (N) and proportion among the grouping variable (e.g. number and percentage of HCWs reporting absent, light, moderate, and heavy for each symptom).

Data management, processing, analysis and visualization were done using R Language and Environment for Statistical Computing (19).

## Results

From the 27^th^ of February until the 8^th^ of April, 2485 NF swabs (some of which repeated) were performed? for HCWs in our two hospitals and the territory, of which 460 NF swabs were done at random. These NF swabs resulted in a total of 185 SARS-CoV-2-positive workers, of which 12 were found through random swabs. The positive rate in non-random samples was around 10%, while the positive rate among randomly sampled HCWs was around 2.6%.

***Table 1*** shows the characteristics of the HCWs included in our study. The study group was made of 143 COVID-19 positive HCWs employed in the different healthcare structures of our TSSC. The data for San Paolo hospital also include the territorial healthcare institutions since their occupational health surveillance is done by the San Paolo hospital. The majority of HCWs were female (57.3%), and the most prevalent job title was nurse (48.3%), followed by medical doctors (26.6%), and assistant nurses (9.8%). Most workers were tested due to a close contact with a positive colleague (49%), followed by worker-initiated testing due to symptoms and unknown contact (total 28%), and a SARS-CoV-2 positive member of the family (9.8%). Close contact with COVID-19 positive patients was the reason for testing in 7.7% of cases HCWs, while 5.6% were tested on random bases. The HCWs under study submitted an average of 9 daily symptoms reports during their quarantine, ranging from 1 to 26.

**Table 1.**
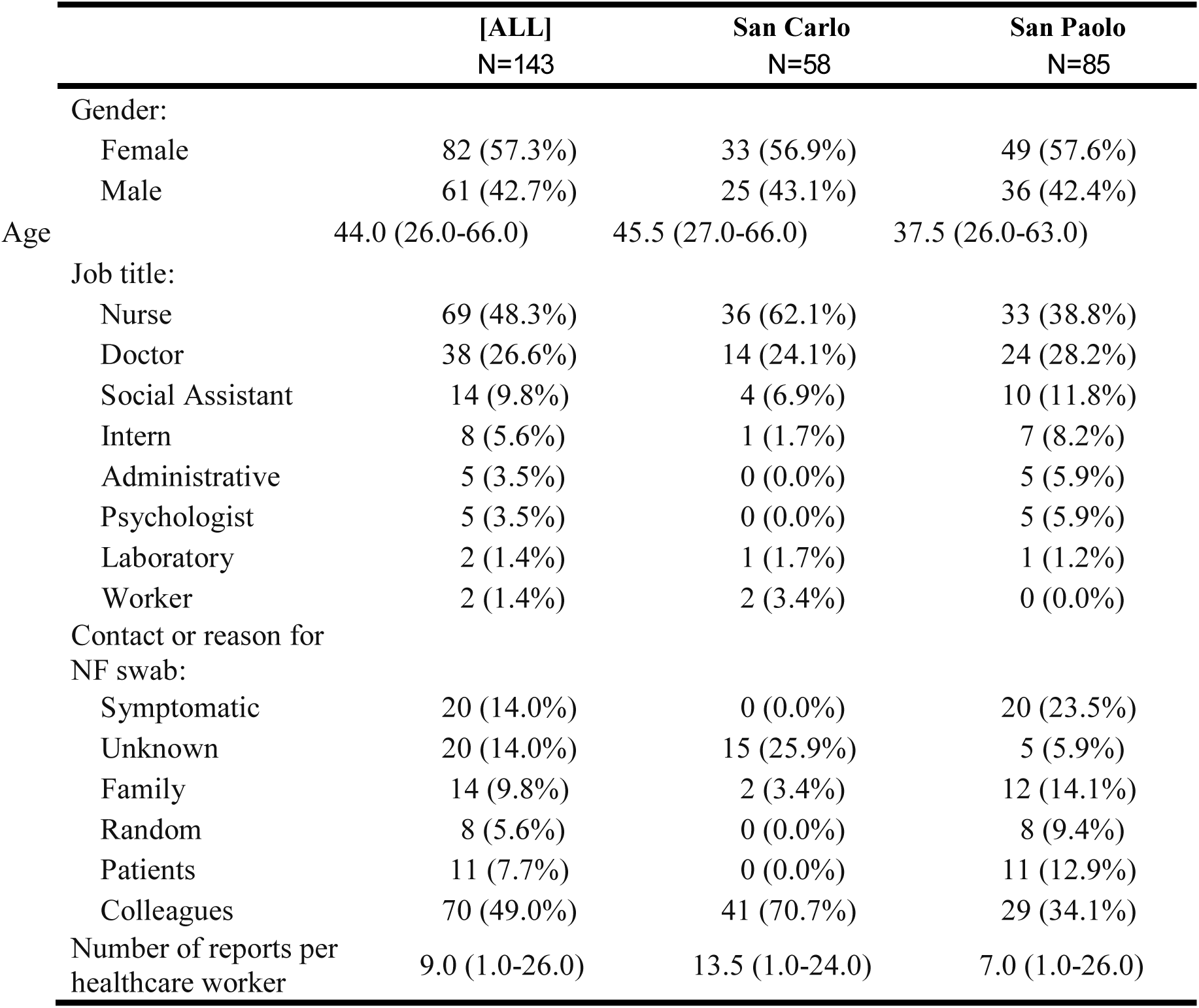
Characteristics of the study population

***Figure 2*** shows timeline of the positive swabs found in HCWs in our study. The first (index) case’s swab was taken on the 25^th^ of February and the result came back on the 27^th^ of February. At the beginning, most individual cases were connected to this case. In the middle weeks of March 2020 the majority of cases (denoted dark red) were connected to other HCWs. The proportion of HCWs infected from close unprotected contacts with colleagues started to decline after the week of 20^th^ of March, and weas replaced by symptomatic HCWs without identified close contact, family-related contacts, and randomly selected for testing HCW.

**Figure 2.**
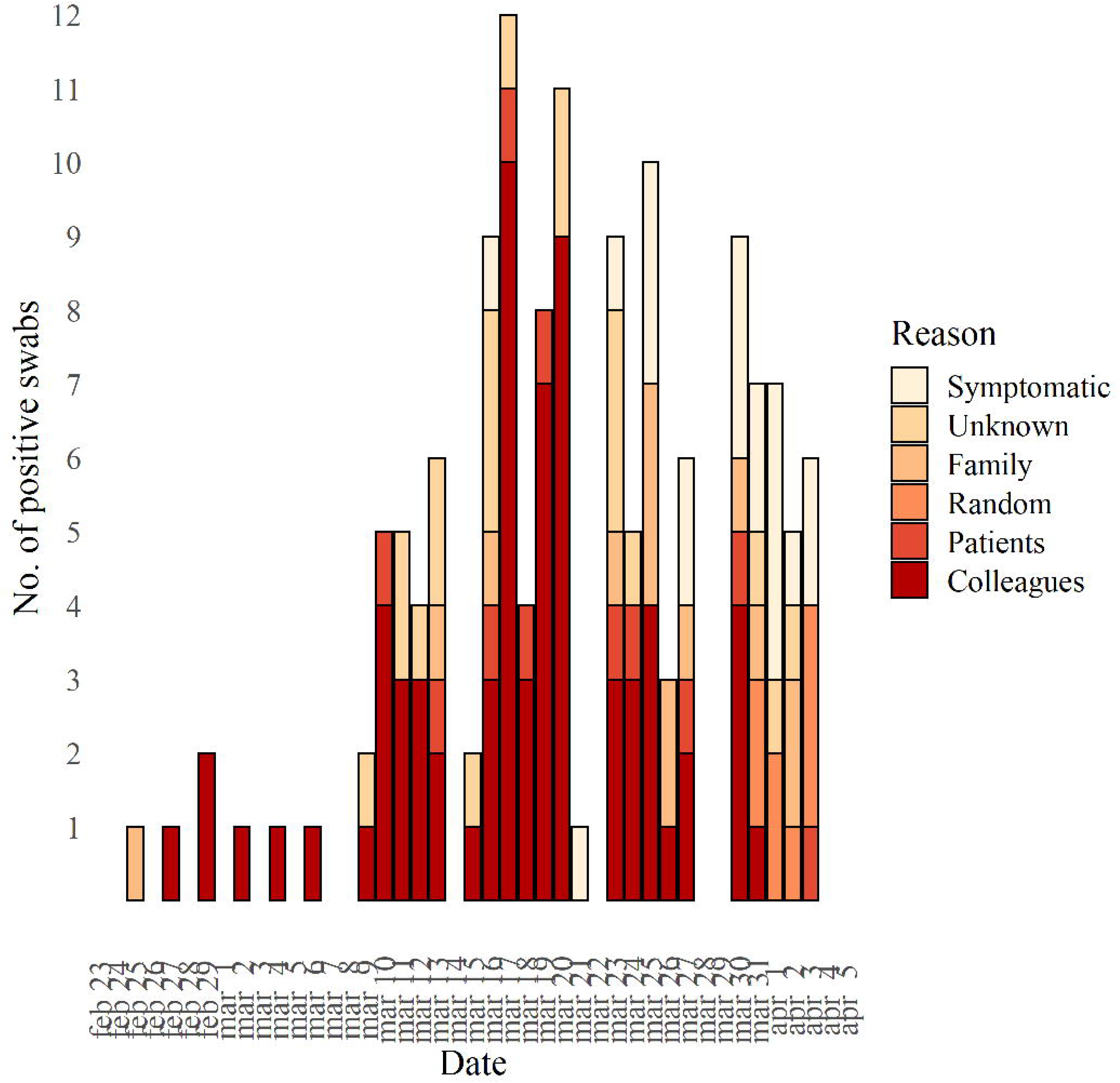
The number of positive swabs since the first (index) case in the hospital.

***Figure 3*** shows the results of contact tracing which started with the first case of a SARS-CoV-2 positive doctor in our study. The doctor had been infected by a family member which later proved to be positive for SARS-CoV-2. Initial contact tracing resulted in 53 “close contacts” inside the hospital, and these 53 swabs resulted in one positive case. From this second positive case other 134 contacts were traced resulting in additional 2 positive HCWs. For each of the new cases another 29 and 20 swabs were performed, resulting in another 2 positive cases on one side, and no positive cases on the other side. This first cluster of cases grew to 7 known cases connected to the index case (data not shown).

**Figure 3.**
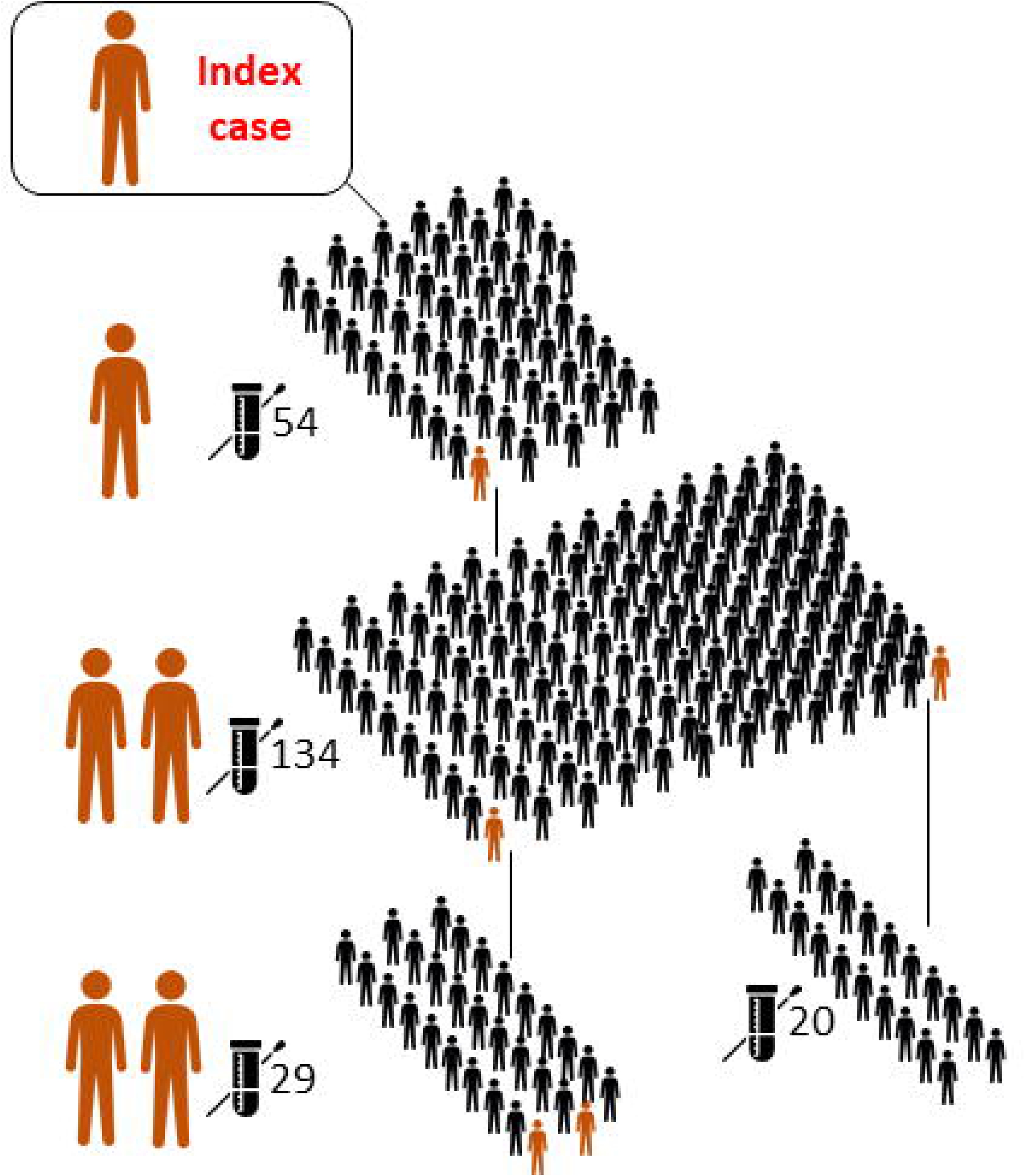
Index case and contact tracing in one of the hospitals.

Most HCWs started submitting daily symptoms reports on the day following the NF swab (66%). The highest percentage of filled reports was noted in the first week, going from 73% on the 2^nd^ day, down to 58% on the 7^th^ day. The gradual drop of daily reports submitted continued in the second week, going from 48% down to 33%. ***Figure 4*** shows the steep rise and gradual decline in the percentage of HCWs submitting the daily health report.

**Figure 4.**
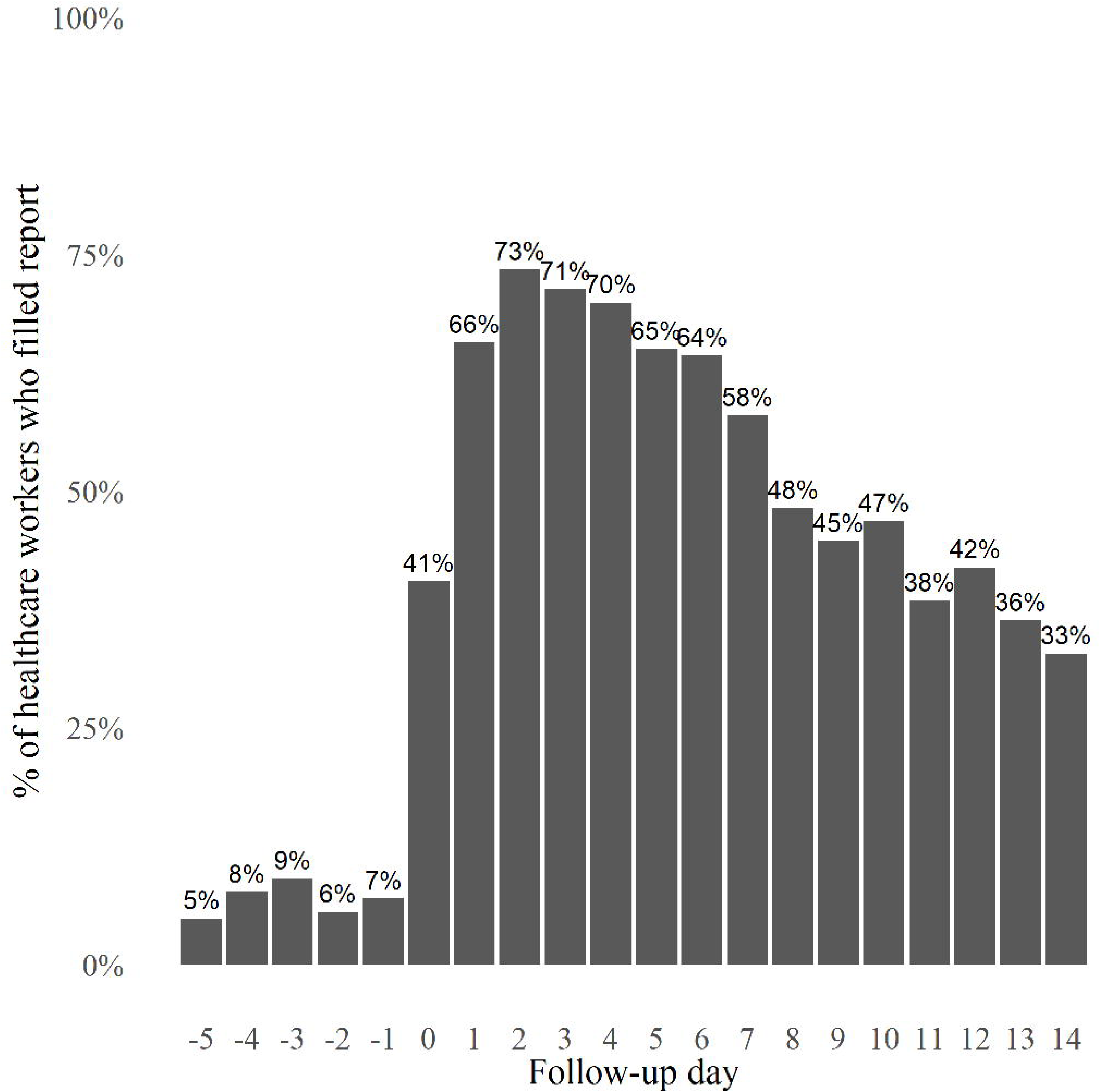
Percentage of HCWs submitting the daily symptoms report relative to the date of the positive NF swab.

***Table 2*** shows the respiratory and other commonly reported symptoms in the day prior to the positive NF swab (***Day* -1**), on the day of the swab (***Day 0***), and in the following day (***Day* 1**). On average, the SARS-CoV-2-infected HCWs were afebrile, with no or light respiratory symptoms. Cough and dyspnoea were absent or light in around 90% of the workers, while general symptoms, such as discomfort, muscle pain, and headache were reported more often, although by no more than 30% of the workers. Most symptoms were absent in more than 50% of the HCWs in the first week of the disease. Anosmia and dysgeusia were the only specific symptoms which were commonly reported, in light, moderate or heavy form by almost 50% of the workers.

**Table 2.** Symptoms of HCWs in the days surrounding the positive NF swab.

|  | **-1** | **0** | **1** |
| --- | --- | --- | --- |
|  | **Day before + NF swab** | **Day of the + NF swab** | **1<sup>st</sup> day after + NF swab** |
|  | **N=10** | **N=58** | **N=94** |
| Morning temperature (°C) | 36.4 (36.0-36.9) | 36.6 (36.0-38.3) | 36.3 (35.2-38.5) |
| Evening temperature (°C) | 36.6 (36.0-37.0) | 36.6 (35.0-38.5) | 36.6 (35.0-38.6) |
| Cough: |  |  |  |
| Absent | 4 (40.0%) | 24 (42.9%) | 43 (45.7%) |
| Light | 3 (30.0%) | 25 (44.6%) | 35 (37.2%) |
| Moderate | 3 (30.0%) | 6 (10.7%) | 14 (14.9%) |
| Heavy | 0 (0.0%) | 1 (1.8%) | 2 (2.1%) |
| Dyspnea: |  |  |  |
| Absent | 9 (90.0%) | 54 (96.4%) | 83 (88.3%) |
| Light | 1 (10.0%) | 2 (3.6%) | 10 (10.6%) |
| Moderate | 0 (0.0%) | 0 (0.0%) | 1 (1.1%) |
| Discomfort: |  |  |  |
| Absent | 5 (50.0%) | 23 (41.1%) | 44 (46.8%) |
| Light | 5 (50.0%) | 17 (30.4%) | 30 (31.9%) |
| Moderate | 0 (0.0%) | 16 (28.6%) | 19 (20.2%) |
| Heavy | 0 (0.0%) | 0 (0.0%) | 1 (1.1%) |
| Muscle pain: |  |  |  |
| Absent | 6 (60.0%) | 27 (48.2%) | 46 (48.9%) |
| Light | 3 (30.0%) | 11 (19.6%) | 23 (24.5%) |
| Moderate | 1 (10.0%) | 16 (28.6%) | 24 (25.5%) |
| Heavy | 0 (0.0%) | 2 (3.6%) | 1 (1.1%) |
| Headache: |  |  |  |
| Absent | 6 (60.0%) | 33 (58.9%) | 56 (59.6%) |
| Light | 3 (30.0%) | 14 (25.0%) | 23 (24.5%) |
| Moderate | 1 (10.0%) | 8 (14.3%) | 12 (12.8%) |
| Heavy | 0 (0.0%) | 1 (1.8%) | 3 (3.2%) |
| Sore throat: |  |  |  |
| Absent | 7 (70.0%) | 34 (60.7%) | 66 (70.2%) |
| Light | 3 (30.0%) | 17 (30.4%) | 19 (20.2%) |
| Moderate | 0 (0.0%) | 5 (8.9%) | 9 (9.6%) |
| Vomiting: |  |  |  |
| Absent | 10 (100.0%) | 54 (96.4%) | 93 (98.9%) |
| Light | 0 (0.0%) | 2 (3.6%) | 1 (1.1%) |
| Anosmia: |  |  |  |
| Absent | 4 (66.7%) | 11 (50.0%) | 24 (55.8%) |
| Light | 1 (16.7%) | 2 (9.1%) | 3 (7.0%) |
| Moderate | 1 (16.7%) | 2 (9.1%) | 7 (16.3%) |
| Heavy | 0 (0.0%) | 7 (31.8%) | 9 (20.9%) |
| Ageusia: |  |  |  |
| Absent | 5 (83.3%) | 12 (54.5%) | 23 (53.5%) |
| Light | 0 (0.0%) | 3 (13.6%) | 6 (14.0%) |
| Moderate | 1 (16.7%) | 2 (9.1%) | 7 (16.3%) |
| Heavy | 0 (0.0%) | 5 (22.7%) | 7 (16.3%) |
| Conjunctival hyperemia: |  |  |  |
| Absent | 6 (100.0%) | 18 (81.8%) | 38 (88.4%) |
| Light | 0 (0.0%) | 3 (13.6%) | 2 (4.7%) |
| Moderate | 0 (0.0%) | 1 (4.5%) | 3 (7.0%) |

***Figure 5*** shows the body temperature of HCWs on the day of the positive NF swab (day 0) and in the 15 days following the swab. ***Figure 6*** shows commonly reported symptoms associated with COVID-19, their intensity, from absent to heavy, as reported by the HCWs during the 15 days following the NF swab. The most common symptoms of a respiratory infection, such as cough, dyspnoea, or sore throat, were present in a heavy or moderate form in less than 15% of the study group in the first days of the infection. Between 70% and 90% of HCWs reported no or mild respiratory symptoms. The only two symptoms reported more commonly as moderate and heavy by between 30% and 40% of HCWs were anosmia and ageusia/dysgeusia. There was a gradual reduction of most symptoms in the second week, with persisting anosmia and dysgeusia in around 30% of HCWs.

**Figure 5.**
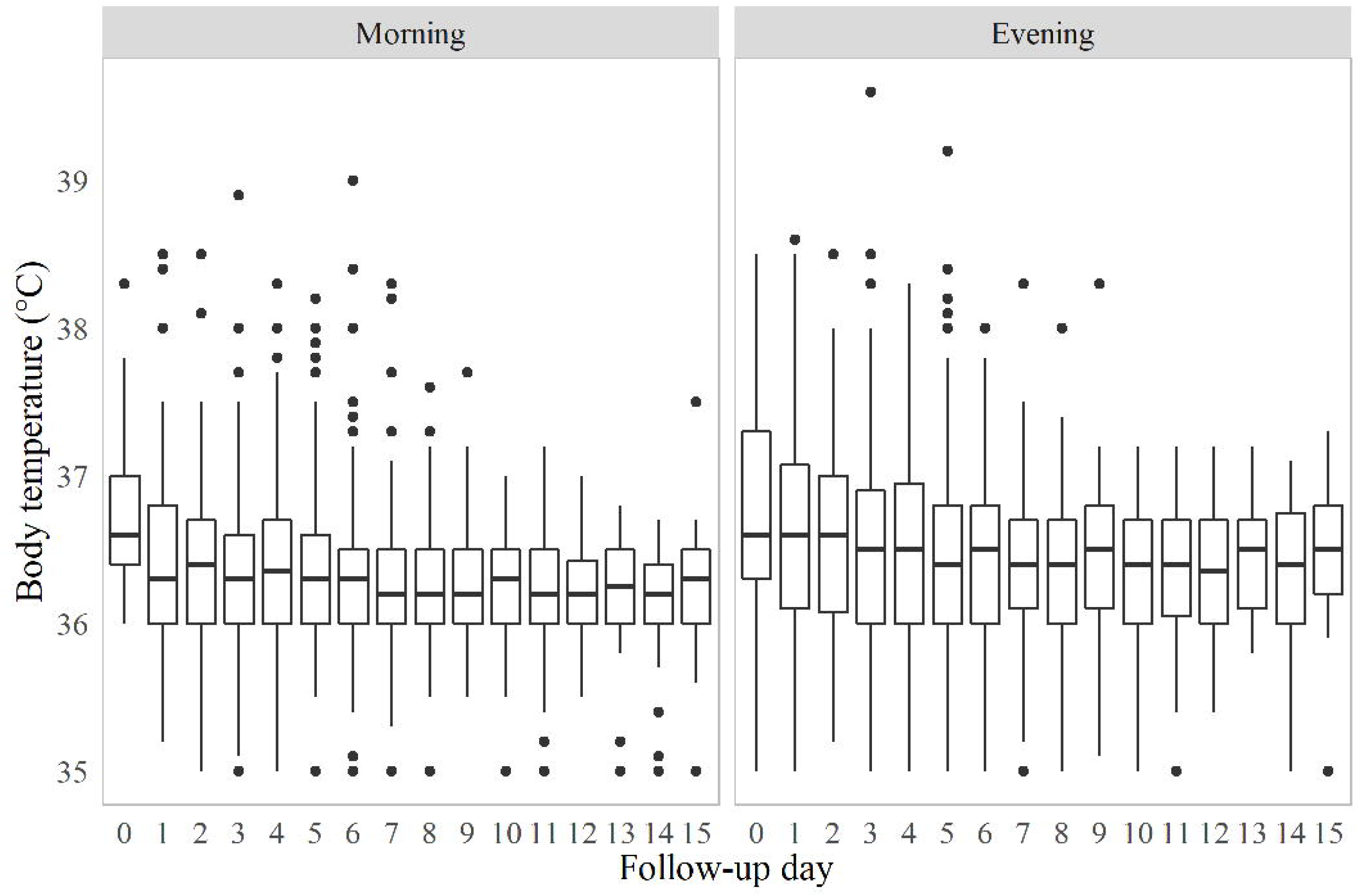
Morning and evening temperature of the SARS-CoV-2 positive HCWs in the first week following the NF swab.

**Figure 6.**
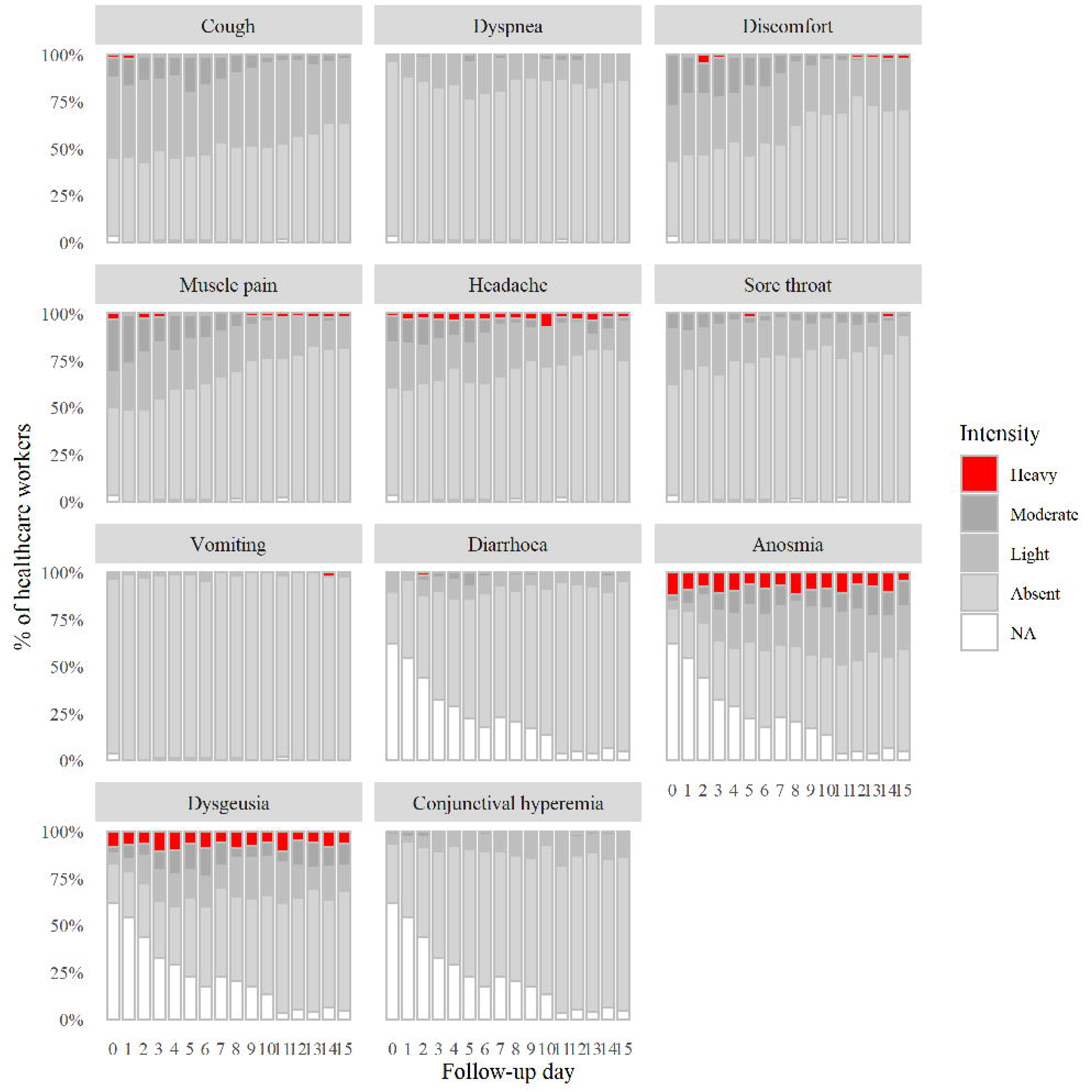
Commonly reported symptoms and signs by HCWs during the 15-day follow up.

For a smaller group of workers who reported their symptoms also in the 5 days leading to the positive NF swab, ***Figure 7*** shows the body temperature and ***Figure 8*** shows other reported symptoms (days −5 to 0). These workers were defined as close contact but were unable to perform a NF swab, so they submitted symptoms reports until the moment of the swab. Although based on a smaller number of reports (see ***Table 2***), most HCWs report normal body temperature or light fever, and those with fevers above 37.5°C or 38°C can be considered outliers. HCWs reported cough (in light and moderate forms), and non-specific symptoms such as discomfort, muscle pain, sore throat, and headache most commonly in the days leading to the positive NF swab. We noted a gradual increase in the number of HCWs reporting anosmia, as well as the gradual worsening of this symptom from light and moderate forms to the heavy form.

**Figure 7.**
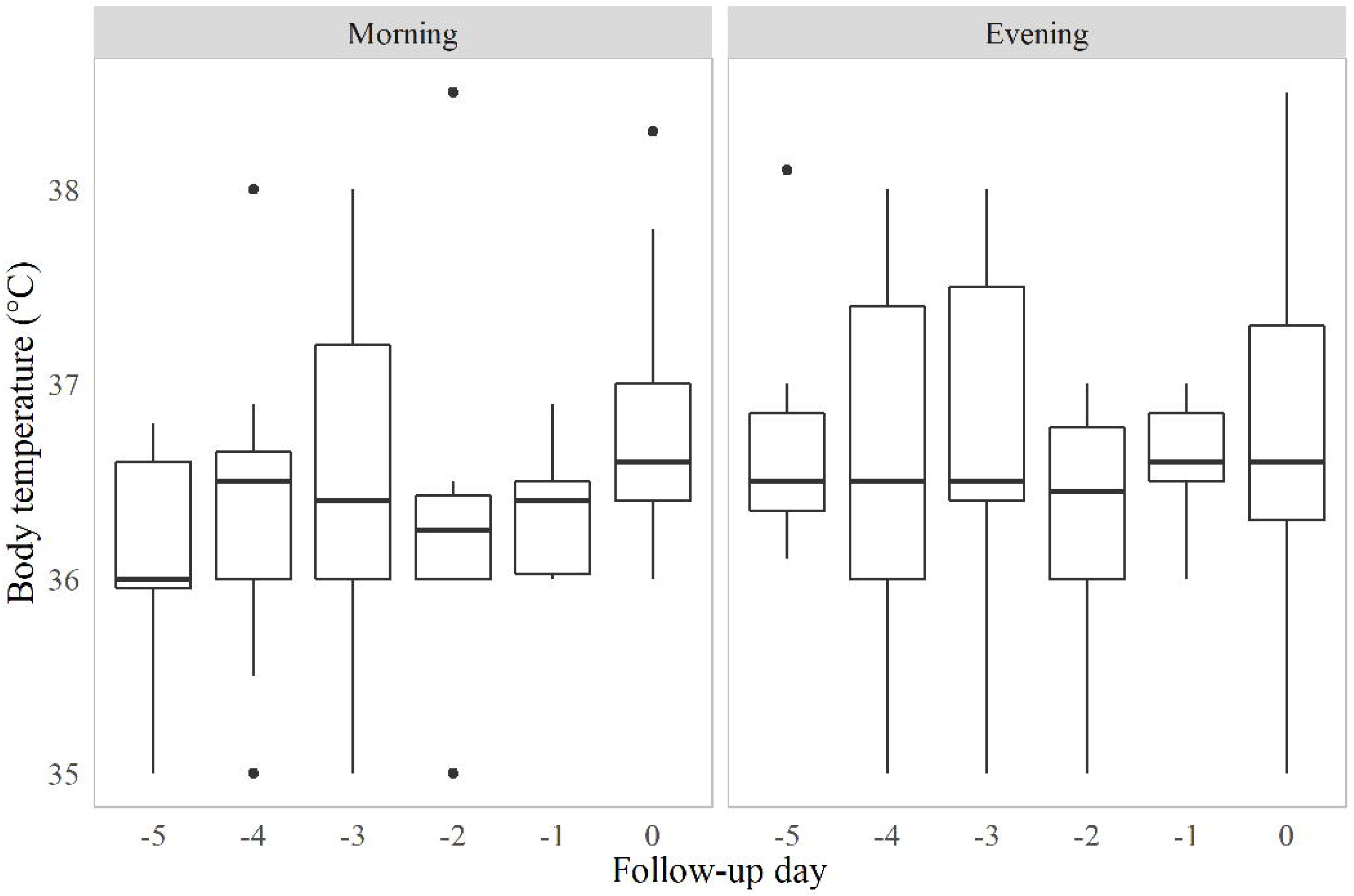
Morning and evening temperature of the SARS-CoV-2 positive HCWs in the 5 days leading to the positive NF swab.

**Figure 8.**
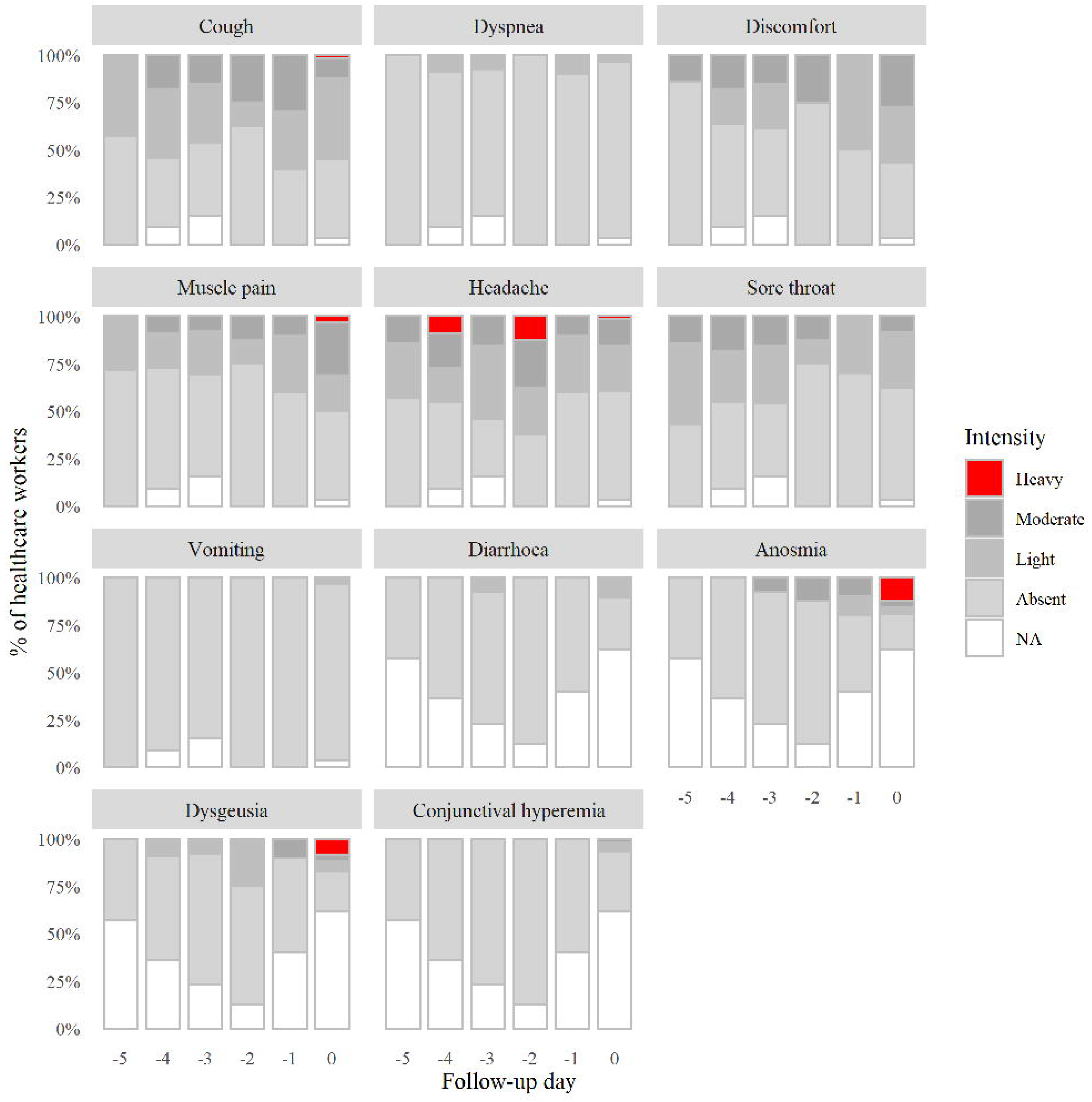
Common symptoms and signs reported by HCWs in the 5 days leading to the positive NF swab.

The median time elapsed from the positive swab to two consecutive negative swabs was 21 days (minimum: 14 days; maximum: 34 days) for San Carlo hospital, and 25 days (minimum: 15 days; maximum: 46 days) for San Paolo hospital (including territorial healthcare institutions). Since the data was not normally distributed, we also report the median values of 20 and 25 in San Carlo and San Paolo hospitals, respectively. 95% of HCWs was considered cured and was able to return to work after 27 days and 36 days in San Carlo and San Paolo hospitals, respectively.

## Discussion

According to the latest data of the Italian Institute of Health (Istituto Superiore Sanita’, April 23^rd^ 2020), there were more than 177 thousand persons positive for SARS-CoV-2 and more than 23 thousand COVID-19 related deaths in Italy. Healthcare workers represent more than 10% of cases (just below 20,000 infected), with a median age of 48 years and 33% male (20). Our results represent one of the first reports on the development of the SARS-CoV-2 epidemic among healthcare workers in large hospital. Our data has shown the number of newly diagnosed SARS-CoV-2-positive HCWs from only one (index case). At the peak of the epidemic in our hospitals and territorial units, up to 12 workers were being diagnosed positive every day, and the source of the infection were other HCWs. Most HCWs had no or only light symptoms which at the beginning never led them or others to doubt they were infective. Common respiratory symptoms, such as fever, cough, dyspnoea, or sore throat were much less reported than recently discovered symptoms such as anosmia and ageusia/dysgeusia. Finally, the time which passes from the positive NF swab to the cessation of symptoms and two consecutive negative NF swabs is more than double of the 14 days initially proposed, which should be taken into account when deciding the duration of the quarantine, even for asymptomatic workers.

The epidemic in our hospitals started with one HCW who was infected outside of the hospital setting. In the first 7 days of the epidemic, we traced more than 250 contacts among HCWs related to this first positive case, which finally resulted in at least 7 positive HCWs. This has been the largest cluster of cases found in our hospital. During the period of around 40 days presented in this report, the Occupational Health Unit of Saints Palo and Carlo hospital performed around 2500 swabs, resulting in 185 positive workers (of which 143 were included in this report), with a much higher percentage of positive cases found among workers identified through contact tracing (~10%) than among randomly sampled (2.6%). The 3^rd^ and 4^th^ weeks of the epidemic in our hospitals were characterized by 5 to 10 cases/day, of which the majority were related to other positive colleagues. Transfer and removal of high-risk workers from high-risk hospital environments, reduction of visiting hours, and avoiding close contacts among patients and colleagues (distance, use of PPD) reduced the potential of SARS-CoV-2 transmission. During the 5^th^ and 6^th^ week of the epidemic in our hospitals, the number of positive HCWs connected to other colleagues declined rapidly, reducing the both the overall number of daily new cases and the size of the clusters surrounding each positive HCW. In the 6^th^ week of tracking, most HCWs who were found positive were tested because they had a positive family member, noticed symptoms, or were tested at random.

The shape of the epidemic curve in our hospital follows the sharp increase of the number of COVID-19 cases in Italy, the Lombardy Region, and Milan. The sharp decline which has followed afterwards in our hospitals does not follow that of Italy as a whole, which was observed several weeks later. We attribute this difference to organizational and protective measures (distancing, use of PPDs for source control) and contact tracing performed by our Occupational Health Unit. Although the number of infected persons in Lombardy has surpassed 60.000, there were only 185 positive HCWs out of 5700 HCWs in our hospitals and the territory, representing just over 3% of workers.

A preliminary report on the epidemic in a large hospital in Madrid, where workers were tested only if presenting at least with mild symptoms, found 791 SARS-CoV-2-positive HCW among around 6800 workers (~11%). The authors found no relation with so-called “high-risk” areas of the hospital, and connect the dynamic of transmission in their hospital to that of the general population (13). Our approach which included contact tracing, source control and testing of asymptomatic workers if they fall under the definition of “close contact” resulted in almost 4 times lower incidence of infection in a setting with more positive cases in the general population. Non-pharmacological measures were underlined as leading to a decline in the effective reproductive number in Wuhan from 3.8 to 0.3, ultimately stopping the epidemic in China (21–23). Similar to the experience in China, our experience underlines the role of asymptomatic patients in driving the epidemic in the hospital but adds workplace transmission between colleagues as a driver of the epidemic among HCWs.

Most HCWs in our study reported no or only mild symptoms of a respiratory infection. In fact, on the day of the positive swab, the median body temperature measured by HCWs was 36.6°C, while cough and dyspnoea were reported as moderate or heavy by only 12.5% and 0% of HCWs, respectively. Unspecific symptoms, such as discomfort, muscle pain, and headache were reported more commonly, but only in around 30% of HCWs. A recent review article on asymptomatic COVID-19 transmission underlined this risk in healthcare setting, although concentrating on the risk arising from asymptomatic patients (24). Our results underline the risk of transmission among asymptomatic HCWs. One of the measures implemented in Lombardy was a mandatory check of the body temperature before the beginning of the work shift for HCWs, and, in case of a temperature above 37.3°C a NF swab is performed and the worker suspended until the results are back (25). Our results show this measure, as any other “symptom-centred” measure, does not guarantee protection of HCWs and patients, and could even result in a false sense of safety in a scenario where up to 90% of cases could be asymptomatic. Most other experiences and opinions regarding the protection of HCWs underline the re-organization of work (e.g. moving triage and pharmacy outside of the hospital), reduction of the density of people (e.g. reducing visiting hours and numbers of visitors), adequate training and use of PPE as the main solutions to protecting HCWs (26–28).

Three additional symptoms drew our attention while working with suspected SARS-CoV-2 positive workers: anosmia, ageusia/dysgeusia, and conjunctival hyperaemia. In fact, since the moment they were added to our daily report in the middle of March, anosmia and ageusia became symptoms most commonly reported as moderate or heavy in SARS-CoV-2 positive HCWs, reported in more than 40% and 30% of cases, respectively. They also represented the most persistent symptoms, as most other symptoms typically connected to respiratory infections exhibited a reduction during the 15-day follow-up. Conjunctival hyperaemia, although noted by our doctors at the hospitals while performing the medical examination and NF swab, was not often reported by HCWs in the daily symptoms report, which might be connected to the fact that it requires an “outside observer” to notice it. It is also important to note that the duration of the infection (time from the positive NF swab to two negative NF swabs) was between 20 and 30 days with 30 days needed for 95% of workers to be considered cured. This information should be considered when deciding whether the quarantine should last for 28 days instead of 14.

A much smaller sample of workers (N=10), commonly found among close contacts but absent from the hospital for other reasons, reported their daily symptoms even in the days leading to the positive NF swab. In the days leading to a positive NF swab, the symptoms and their intensity were similar to those reported during the follow-up period. Cough and non-specific symptoms were reported more commonly, fever and other respiratory symptoms rarely, and we noted a gradual increase in anosmia and ageusia leading to the day of the positive NF swab. This is the first report of symptoms in the pre-swab period of SARS-CoV-2 positive patients.

Our study was based on the data collected to monitor workers health status while quarantined, the filling of the daily symptoms report was voluntary, and the report itself was adapted several times to answer the needs and field situation, which introduce bias in the presented results. Nevertheless, our report is based on a relatively high response rate of around 70% in the first week of follow-up. A limitation is the fact that 42 HCWs never filled in even one daily symptoms report. In a telephone survey currently in progress, the most common reasons for not filling the online symptoms report was the lack of a smartphone, computer or internet at home, lack of experience with online forms (“not being technological enough”), being diagnosed in the week prior to the implementation of the online report, and taking care of a sick family member (“lack of time for the reports”). Five HCWs did not fill the report because they were hospitalized. Their reports would certainly differ from the rest of our workers with a mild clinical picture, but heir percentage (less than 5% of 185), similar to that in Madrid where 29 out of 791 required hospitalization, leads us to believe that the data presented are representative of the majority SARS-CoV-2-positive HCWs and the selection bias in our report is negligible.

Future studies should analyse in more detail the circumstances surrounding the infection of HCWs, symptoms, and the overall outcome of their disease. New symptoms, such as anosmia and dysgeusia which were frequently reported by our HCWs could help clinicians arrive to a diagnosis sooner and reduce the time available for worker-to-worker and worker-to-patient transfer of SARS-CoV-2. Another step forward will be understanding whether HCWs have developed specific immunity, even among those with negative swab results. This could help us understand whether the presence of specific IgG could be an expression of effective immunity, as well as whether it is temporary or permanent. Finally, the lessons learned from controlling the SARS-CoV-2 epidemic among HCWs could be applied to other occupations/sectors at risk, such as transport workers, services and sales workers, and public safety workers.

## Conclusions

HCWs represent one of the most important resources in the fight against COVID-19, but they are also one of the most vulnerable groups which is commonly infected. Our study has shown that at the peak of the epidemic in Italy the reorganization of work, physical distancing, use of PPDs (even for source control), and contact tracing and testing of asymptomatic HCWs were able to stop the worker-to-worker infection in the hospital and reduce the overall incidence of infection among HCWs. Most HCWs were asymptomatic or with mild symptoms, which means they would most likely go undetected by symptom-centred preventive strategies. Most reported specific symptoms were anosmia and ageusia, and their role in arriving to a clinical diagnosis sooner should be further confirmed. Finally, our data suggest that the duration of the infection is longer than previously anticipated, and that a patient should not be considered recovered only 14 days after the positive swab. Readmitting such a patient or worker into the social or work life without a further swab assessment might create additional risk.

## Data Availability

Data is still being updated and elaborated and is currently not available.

## Acknowledgements

The Authors would like to acknowledge the kind help and support from our dear colleagues Rosamaria Bentoglio, Donatella Visentin and all workers of the Occupational Health Unit of the Saints Paolo and Carlo Hospital of Milan without whose selfless effort this work would not be possible.

## Authors Contributions

SMR: literature search, figures, study design, data collection, data analysis, data interpretation, original manuscript writing, manuscript revision; FM: literature search, figures, study design, data interpretation, original manuscript writing, manuscript revision; EC: study design, data collection, manuscript revision; SF: study design, data collection, manuscript revision; AL: study design, data collection, manuscript revision; IB: study design, data collection, manuscript revision; SV study design, data collection, manuscript revision; AA: study design, data collection, manuscript revision; RB: study design, data collection, manuscript revision; LB: study design, data collection, manuscript revision; LN: study design, data collection, manuscript revision; AZ: study design, data collection, manuscript revision; VO: study design, data collection, manuscript revision; GO: study design, data collection, manuscript revision, laboratory analyses; CC: study design, data collection, manuscript revision, supervision, data interpretation, manuscript revision.

## Funding statement

This study received no funding. All Authors are employed in the Institution (see affiliation) where the study was conducted.

The corresponding authors had full access to all the data in the study and had final responsibility for the decision to submit the publication.

